# Genomic insights into extrapulmonary tuberculosis reveal enrichment of lineage 2 with high drug resistance in specific clinical phenotypes

**DOI:** 10.1101/2025.08.05.25332932

**Authors:** Kusum Sharma, Aravind Bhandari, Gowrang Kasaba Manjunath, Mona Rastogi, Aishwarya S. Babu, Parthasarathy Ponnusamy, Shruthi Vasanthaiah, Karthick Vasudevan, Priyanka Yadav, John George, Abhishek Kumar, Jyoti Sharma, Aman Sharma, Manish Modi, Amod Gupta, Ramandeep Singh, Saroj. K. Sinha, M.S. Dhillon, Megha Sharma, Vishal Sharma, Ritambhra Nada, Navneet Sharma, Akhilesh Pandey, Renu Verma

## Abstract

**Background:** Certain *M. tuberculosis* lineages may preferentially infect extrapulmonary sites, potentially influencing clinical outcomes. Poor drug penetration at these sites may cause treatment failure, even in the absence of resistance. Timely access to resistance profiles and lineage information may support more effective, personalized treatment for extrapulmonary tuberculosis (EPTB).

**Methods:** We sequenced 117 *M. tuberculosis* culture isolates from seven EPTB sites at PGIMER, India. This included cerebrospinal fluid (CSF, n = 40), pus (n = 35), fine-needle aspiration cytology (FNAC, n = 22), tissue (n = 8), ileocaecal biopsy (n = 6), synovial fluid (n = 5), and vitreous fluid (n = 1). Phenotypic drug susceptibility testing (pDST) against 12 drugs was performed using the MYCOTBI sensititre assay. Whole genome sequencing (WGS) on Illumina XTen platform and analyzed using in-house pipelines.

**Results:** Drug resistance was identified in 23.9% (28/117) of isolates by pDST and 29.9% (35/117) by WGS, with 93.0% concordance. WGS detected additional resistant cases missed by MYCOTBI (p = 0.039). Resistance was significantly associated with sample type (p = 0.0446), highest in CSF (16/40, 40.0%). Lineage 2 had the highest resistance (13/24, 54.1%), primarily from CSF (9/13, 69.2%). Mixed infections were observed in 6.8% (8/117) of isolates, mostly involving lineages 2 and 3. Heteroresistance was more common in mixed infections (p = 0.0139).

**Conclusions:** WGS reliably detected resistance to anti-TB drugs, along with lineage and mixed infections. This approach can be applied in culture-free targeted sequencing for rapid detection of resistance and lineage, enabling personalized treatment regimens and improved outcomes in EPTB.

## Introduction

EPTB accounts for 15–20% of the global TB burden [1] and is associated with high mortality, poor treatment outcomes, and increased relapse rates [2–5]. Recent studies have reported rising drug resistance among EPTB patients, complicating diagnosis and disease management [6,7]. Drug resistance in EPTB ranges from 10–20%, with up to 40% resistance reported against first-line anti-TB drugs [8].

Despite these challenges, current guidelines largely recommend uniform treatment regimens for pulmonary TB and EPTB [9]. However, anti-TB drugs show lower penetration at certain extrapulmonary sites, such as CSF, leading to subtherapeutic levels and possible treatment failure [10]. In such cases, poor drug exposure rather than microbial resistance may contribute to adverse outcomes. Dose adjustments based on the clinical phenotype and resistance profile may improve outcomes [11].

Compounding these challenges, clinicians often lack complete resistance profiles to distinguish between pharmacokinetic failure and true resistance due to challenges in detecting drug resistance in EPTB samples. In addition, *M. tuberculosis* lineage influences disease severity, transmission dynamics, and resistance development [12]. Some lineages exhibit higher virulence, increased drug resistance, and stronger associations with specific anatomical sites in pulmonary TB [13], yet their role in EPTB remains underexplored.

Diagnosing drug resistance in EPTB is further complicated by its paucibacillary nature and low culture positivity [6]. Resistance is frequently suspected only after clinical non-response [14]. The WHO recommends molecular assays like GeneXpert MTB/RIF and GeneXpert MTB/RIF Ultra for rapid detection of *M. tuberculosis* and rifampicin resistance, though these tools detect limited mutations [2]. Studies have identified resistance to both first- and second-line drugs in EPTB [15,16].

Targeted sequencing platforms, informed by the WHO mutation catalogue, enable rapid, culture-free detection of resistance mutations and lineage assignment [17,18]. However, these panels are primarily developed using data from pulmonary TB, limiting their generalizability to EPTB [19]. Furthermore, phenotypic data for drugs like ofloxacin, cycloserine, para-aminosalicylic acid (PAS), and rifabutin are not included in the current WHO catalogue, despite evidence of resistance-linked mutations [20–22].

In this study, we performed WGS of *M. tuberculosis* isolates from seven extrapulmonary sites and compared genotypic resistance with MYCOTBI phenotypic DST across twelve anti-TB drugs. We also examined heteroresistance, mixed infections, and associations between lineage, drug resistance, and clinical phenotype.

## METHODS

### Patient enrollment and ethical statement

We used *M. tuberculosis* positive culture isolates from extrapulmonary sites collected between 2014-2020 at the Department of Medical Microbiology, Postgraduate Institute of Medical Education and Research (PGIMER), India. The study was approved by the PGIMER Institutional Ethics Committee (Reference No. NK/1691/Res/3286). All participants were aged 18 years or older and provided written consent before the initiation of the study.

### Sample processing and DNA extraction

A total of 117 *M. tuberculosis* positive cultures were collected from seven extrapulmonary sites. These included CSF (n = 40), pus (n = 35), fine-needle aspiration cytology (FNAC) (n = 22), tissue (n = 8), ileocaecal biopsy (ICBx) (n = 6), synovial fluid (SF) (n= 5) and vitreous fluid (VF) (n =1). All the isolates were processed as per the standard protocol described previously [23]. The samples were inoculated to Mycobacteria Growth Indicator Tube (MGIT) and solid culture Lowenstein-Jensen (LJ) slants (Difco Lowenstein Medium Base No. 244420) and incubated at 37°C for eight weeks. The cultures were subsequently tested using commercially available SD BIOLINE TB Ag MPT64 rapid kit to rule out non-tuberculous mycobacteria. DNA was extracted using QIAamp DNA Mini Kit, according to the manufacturer’s instructions, eluted in 50µl nuclease-free water and stored at -20°C until use.

### Sensititre MYCOTB assay

We performed minimum inhibitory concentration (MIC) based phenotypic DST using MYCOTBI sensititre plates (Sensititre MYCOTB assay, Thermo Fisher Scientific, USA) on all culture-positive samples (n = 117) according to the manufacturer’s instructions. The MYCOTBI plates are pre-coated with twelve standardized drug concentrations, including isoniazid, rifampicin, ethambutol, streptomycin, moxifloxacin, ofloxacin, cycloserine, PAS, rifabutin, amikacin, kanamycin, and ethionamide [24].

For MYCOTBI testing, mycobacterial strains isolated from extrapulmonary regions were subcultured onto LJ slants (Difco Lowenstein Medium Base No. 244420). Colonies with bacterial growth were added to 200 μl of saline-Tween 80 solution (Trek Diagnostic Systems) containing 0.5 mm silica beads and vortexed for 30 seconds. The colony suspension was allowed to settle for 10 minutes, and the turbidity of the sample was adjusted to match the 0.5 McFarland standard. A total of 100 μl of the adjusted saline suspension was transferred to 11 ml of Middlebrook 7H9 broth containing oleic acid-albumin-dextrose-catalase (Trek Diagnostic Systems) and mixed thoroughly. Then, 100 μl of the suspension was inoculated into each well of the MYCOTBI plate. The plates were sealed and incubated at 37°C in a 5% CO atmosphere. Plates were monitored on days 7, 14, 21, and 28 post-inoculation using a manual plate reader with data management software (Vizion System, TREK Diagnostics). If the MIC value of a drug was higher than its critical concentration, the strain was classified as resistant; if it was equal to or below the critical concentration, the strain was classified as sensitive. Each MYCOTBI plate was independently read, interpreted, and recorded by two readers to ensure agreement. For phenotype-to-genotype comparisons of drug resistance, the readings from day 28 were considered final.

### Library preparation

Genomic DNA was quantified using the Qubit fluorometric method. DNA samples were prepared for library construction using the Twist EF Library Prep Kit for Illumina (Cat#100572). Initially, DNA was fragmented to the desired size, followed by end-repair and 3’ mono-adenylation in a single enzymatic reaction. Adapters were then ligated using a T4 DNA ligase-based reaction. Post-ligation, the libraries were PCR-amplified with unique barcoded primers (Twist Unique Dual Index Primer Set A, Cat#101308) for sample multiplexing. Fragment distribution was assessed using the Agilent 5300 Fragment Analyzer System. The clustered libraries were sequenced on the Illumina HiSeq XTen platform to generate 150 bp paired-end reads.

### Variant mapping, annotation and phylogenetic analysis

We used Snippy [25] for whole-genome alignment to the *M. tuberculosis* reference genome H37Rv (Acc: NC000962) and variant calling. Snippy is a freely available Perl-based tool capable of identifying both single nucleotide polymorphisms (SNPs) and indels. It leverages BWA-MEM [26] and FreeBayes [27]. We ran Snippy with default parameters, which have previously been shown to produce high-quality variant calls. Variants were annotated using SnpEff [28] with the H37Rv reference genome. Snippy-core was employed to generate core-alignment files. Gubbins v2.4.1 [29] was used to produce an alignment file with recombinant sites masked. A maximum-likelihood phylogenetic tree based on the core SNPs was constructed using IQ-TREE2 [30], with 1000 ultrafast bootstrap iterations and the GTR model. The generated phylogenetic tree was visualized and annotated using iTOL [31].

### Drug Resistance and lineage identification

We used TBProfiler v6.5.0 to detect drug resistance and identify lineages [32]. TBProfiler aligns reads to the H37Rv reference genome using Bowtie2, BWA, or Minimap2, and then calls variants with BCFtools. After variant calling, the collate function extracts drug-resistance mutations and lineage information using a database of drug resistance mutations and lineage markers hosted on the TBDB repository [32]. Additionally, the identified mutations associated with drug resistance were compared against the WHO catalogue [33] to distinguish between mutations linked to drug resistance and those not associated with it.

### Heteroresistance detection

We used an inhouse customized Python scripts [34] to detect heteroresistance in *M. tuberculosis*. Initially, we curated a database of mutations associated with antibiotic resistance [35]. Using the samtools mpileup command, we generated pileup files for specific genomic regions of interest from a sorted BAM file aligned to the reference genome of *M. tuberculosis H37Rv*. We then parsed these pileup files to calculate nucleotide frequencies at the mutation sites. Further, we processed each mutation entry by extracting the reference and mutant nucleotide frequencies, as well as the total read counts, to determine the presence and extent of heteroresistance. We used a minimum 10 reads cutoff to consider a mutation as true positive for heteroresistance detection. The calculated frequencies and read counts were subsequently compiled into an Excel report, sorted by mutant frequency to highlight significant mutations.

### Data availability

WGS data were deposited in the National Center for Biotechnology Information (NCBI) Sequence Read Archive (SRA) database with accession PRJNA1273668 (https://www.ncbi.nlm.nih.gov/sra/PRJNA1273668)

### Statistical analysis

A Chi-square test was conducted to assess the presence of a significant association between sample type and drug resistance. Only sample types with a sample size of 10 or greater were included in the analysis. We also performed Chi-square test to determine if there was any significant correlation between sample type, lineage, and drug resistance. We performed McNemar’s test to compare the diagnostic performance of MYCOTBI phenotypic drug susceptibility testing with whole genome sequencing, considering WGS as the reference standard. All data analysis were performed in R.

## RESULTS

### Study participants and sample characteristics

Of the 117 individuals with active EPTB whose positive culture isolates were used for the study, 58% (68/117) were male (median age = 37 years; IQR = 30–48). The median age of female participants was 33 years (IQR = 24.5–55.5). Samples were classified based on the site of infection, including CSF (40/117, 34.2%), pus (35/117, 29.9%), FNAC (22/117, 18.8%), tissue (8/117, 6.8%), ICBx (6/117, 5.1%), SF (5/117, 4.3%), and VF (1/117, 0.9%). The clinical and microbiological characteristics of the study cohort are summarized in **Table 1**.

**Table 1:** Clinical and microbiological characteristics of study cohort.

| **Clinical characteristic** |  |
| --- | --- |
| Total no of patients (n) | 117 |
| Male, n (%) | 68 (58.1%) |
| Male, age in years, median (IQR) | 37 (30.0-48.0) |
| Female, age in years, median (IQR) | 33 (24.5-55.5) |
| **Sample type** |  |
| Cerebrospinal fluid (CSF), n (%) | 40 (34.2%) |
| Pus, n (%) | 35 (29.9%) |
| Fine needle aspiration cytology (FNAC), n (%) | 22 (18.8%) |
| Tissue, n (%) | 8 (6.8%) |
| Ileocecal biopsy (ICBx), n (%) | 6 (5.1%) |
| Synovial fluid (SF), n (%) | 5 (4.3%) |
| Vitreous fluid (VF), n (%) | 1 (0.9%) |
| **MYOCTBI profile<sup>1</sup>** |  |
| DS-TB<sup>2</sup>, n (%) | 89 (76.1%) |
| HR-TB<sup>3</sup>, n (%) | 17 (14.5%) |
| RR-TB<sup>4</sup>, n (%) | 2 (1.7%) |
| MDR-TB<sup>5</sup>, n (%) | 1 (0.9%) |
| Pre-XDR TB<sup>6</sup>, n (%) | 8 (6.8%) |

MYCOTBI profile was classified according to the latest WHO guidelines<sup>1</sup>, Drug sensitive tuberculosis<sup>2</sup>, Isoniazid-resistant tuberculosis<sup>3</sup>, Rifampicin-resistant tuberculosis<sup>4</sup>, Multidrug-resistant tuberculosis<sup>5</sup>, Pre-extensively drug-resistant tuberculosis<sup>6</sup>, IQR; Interquartile range

### MYCOTBI phenotypic drug susceptibility

Of the seven different sample types, resistance to at least one drug tested in the MYCOTBI phenotypic drug susceptibility assay was observed in 28/117 (23.9%) *M. tuberculosis* isolates. These included samples from four sites, CSF (16/28, 57.1%), pus (8/28, 28.5%), synovial fluid (SF, 1/28, 3.5%), and FNAC (3/28, 10.7%). Based on MYCOTBI phenotypic DST, samples were identified as isoniazid-resistant TB (HR-TB, 17/117, 14.5%), rifampicin-resistant TB (RR-TB, 2/117, 1.7%), multidrug-resistant TB (MDR-TB, 1/117, 0.9%), and pre-extensively drug-resistant TB (Pre-XDR-TB, 8/117, 6.8%). We observed high levels of drug resistance to first-line anti-TB drugs. Isoniazid mono-resistance was the most prevalent (17/28, 60.7%), followed by resistance to ethambutol (13/117, 11.1%), rifampicin (11/117, 9.4%), and streptomycin (11/117, 9.4%). No resistance was detected to ofloxacin, cycloserine, PAS, or rifabutin in phenotypic drug susceptibility testing. (**Supplementary table 1**).

### Whole genome sequencing and genotypic drug resistance

All samples (n = 117) met the minimum quality criteria (Phred score ≥ 20, coverage depth ≥ 50x). The median H37Rv genome coverage across all samples was 99.1% (IQR = 99.3–99.8). The overall median coverage depth was 201x (IQR = 122.0–339.5). A summary of sequencing statistics is provided in **Table 2A**. We observed higher median coverage depth in CSF (median 257.2x) and pus samples (median 320x) compared to other sample types (**Figure 1**).

**Table 2A:** Sequencing statistics of 117 *M. tuberculosis* genomes from EPTB samples.

| **Sample type**<br>**(n)** | **Mean baseQ,**<br>**median (IQR)** | **Reads**<br>**mapped to**<br>**H37Rv (n)** | **H37Rv genome**<br>**coverage (%)** | **H37Rv genome**<br>**coverage depth**<br>**(x)** |
| --- | --- | --- | --- | --- |
| CSF (40) | 37.6 (36.8-38.5) | 4,958,397 | 99 (98.8-99.2) | 257 (144.3-339.3) |
| Pus (35) | 37.4 (36.8-38.3) | 6107067 | 99.2 (98.9-99.3) | 320 (199.5-417.5) |
| FNAC (22) | 37.3 (37-37.4) | 3,408,569 | 98.9 (98.5-99.1) | 143 (111.8-176.8) |
| Tissue (8) | 38.5 (38.4-38.5) | 2,244,612 | 99.4 (99.3-99.4) | 143 (78-296.8) |
| ICBx (6) | 36.9 (36.8-37.1) | 3447245 | 99.4 (99.2-99.5) | 141 (124.2-158.2) |
| SF (5) | 38.5 (37.3-38.8) | 2,707,173 | 99.4 (99-99.5) | 158 (101-230) |
| VF (1) | 36.9 | 3,284,058 | 98.9 | 132 |
| Total samples<br>(117) | 37.3 (36.8-38.4) | 3,992,798 | 99.1 (98.8-99.3) | 201 (122-339) |

CSF- Cerebrospinal fluid, FNAC- Fine-needle aspiration cytology, ICBx- Ileocaecal biopsy, SF- Synovial fluid, VF- Vitreous fluid, IQR- Interquartile range

**Figure 1:**
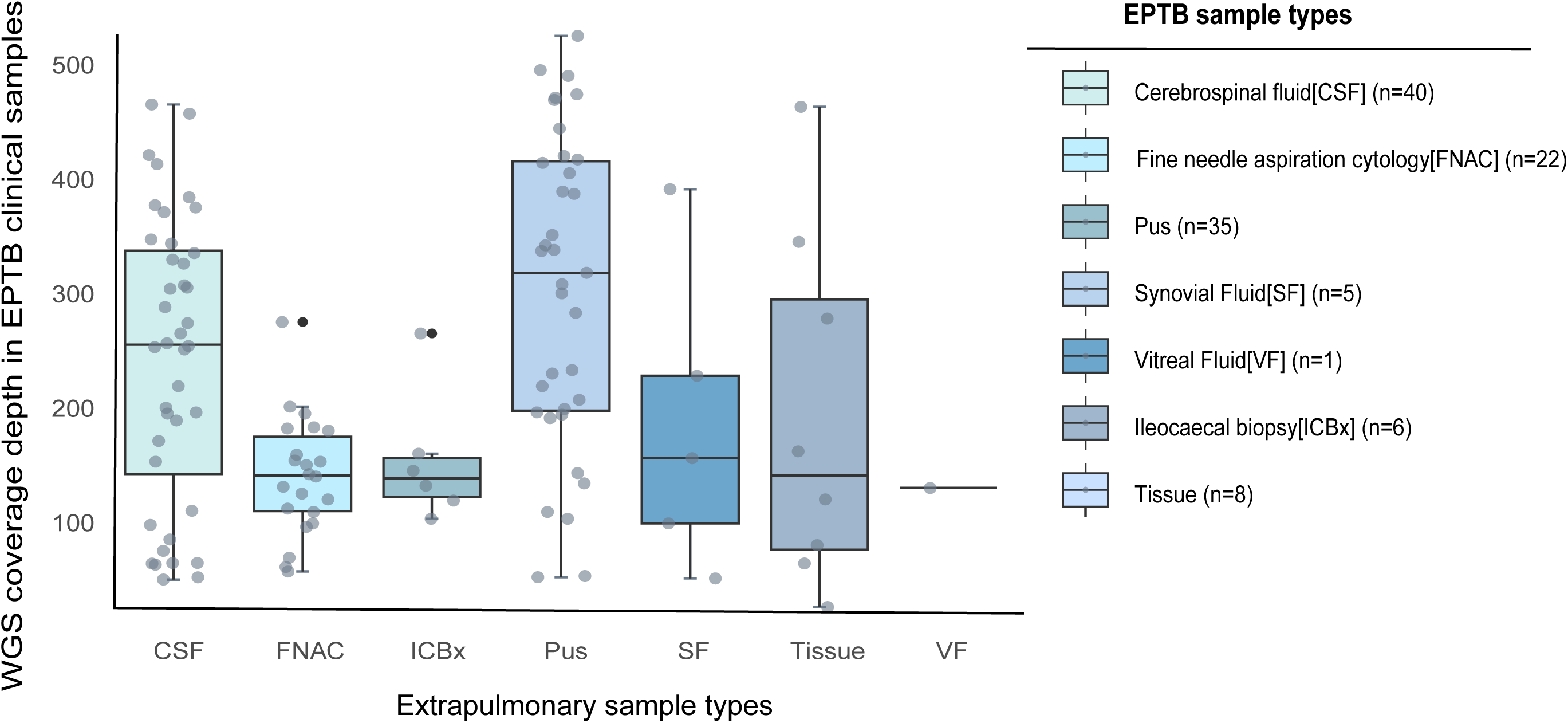
WGS coverage depth in EPTB clinical samples. The samples were categorized based on their site of infection and categorized into seven types - cerebrospinal fluid (CSF), fine needle aspiration cytology (FNAC), ileocecal biopsy (ICBx), pus, synovial fluid (SF), tissue and vitreous fluid (VF). Gray dots represent the individual sample in each category. The median genome coverage of the H37Rv strain across all samples was 99.1% (IQR = 99.3–99.8). The overall median coverage depth was 201x (IQR = 122.0–339.5). We observed higher median coverage depth in CSF (median 257.2x) and pus samples (median 320x) compared to other sample types.

We compared the variant calling data from WGS with the most recent version of the WHO mutation catalogue to identify drug resistance-associated mutations [33]. Mutation profiles for four drugs included in the MYCOTBI sensititre panel namely ofloxacin, cycloserine, PAS, and rifabutin are not present in the WHO mutation catalogue. However, substantial literature evidence supports the existence of drug resistance-associated mutations for these drugs [20–22]. These mutations are incorporated into the widely used TBProfiler platform [32]. We used the mutation profiles derived from TBProfiler for these drugs to assess genotype-to-phenotype concordance. Based on WGS analysis, 36/117 (30.7%) samples exhibited mutations associated with at least one drug included in the MYCOTBI test panel (**Figure 2A, Supplementary Table 2**). Seventeen unique mutations were identified in eight genes (*embA, embB, fabG1, katG, gyrA, pncA, rpoB, rrs*). The *katG* p.Ser315Thr mutation, which confers high-level resistance to isoniazid, was detected in 21/117 (17.9%) samples. Rifampicin resistance associated mutations were detected in 18/117 (15.3%) samples. These mutations are included in the GeneXpert MTB/RIF Ultra point-of-care test and are known to confer high-level rifampicin resistance [36]. The *rpoB* p.Ser450Leu mutation was the most prevalent, detected in 11/18 samples (61.1%), followed by *rpoB* p.His445Asn in 4/18 samples (22.2%), and *rpoB* p.Leu452Pro in 2/18 samples (11.1%). These mutations occur within the rifampicin resistance-determining region (RRDR) of the *rpoB* gene and are associated with high-level rifampicin resistance. Of the four drugs for which mutation profiles are not yet fully incorporated in the current WHO mutation catalogue, we detected mutations associated with ofloxacin resistance in 12 samples and rifabutin resistance in 18 out of 117 samples. According to WHO’s 2021 catalogue of mutations in *M. tuberculosis,* the p.Asp435Val mutation is classified as associated with rifampicin resistance but does not always confer resistance to rifabutin. The distribution of mutations across various samples is shown in **Table 2B**.

**Figure 2:**
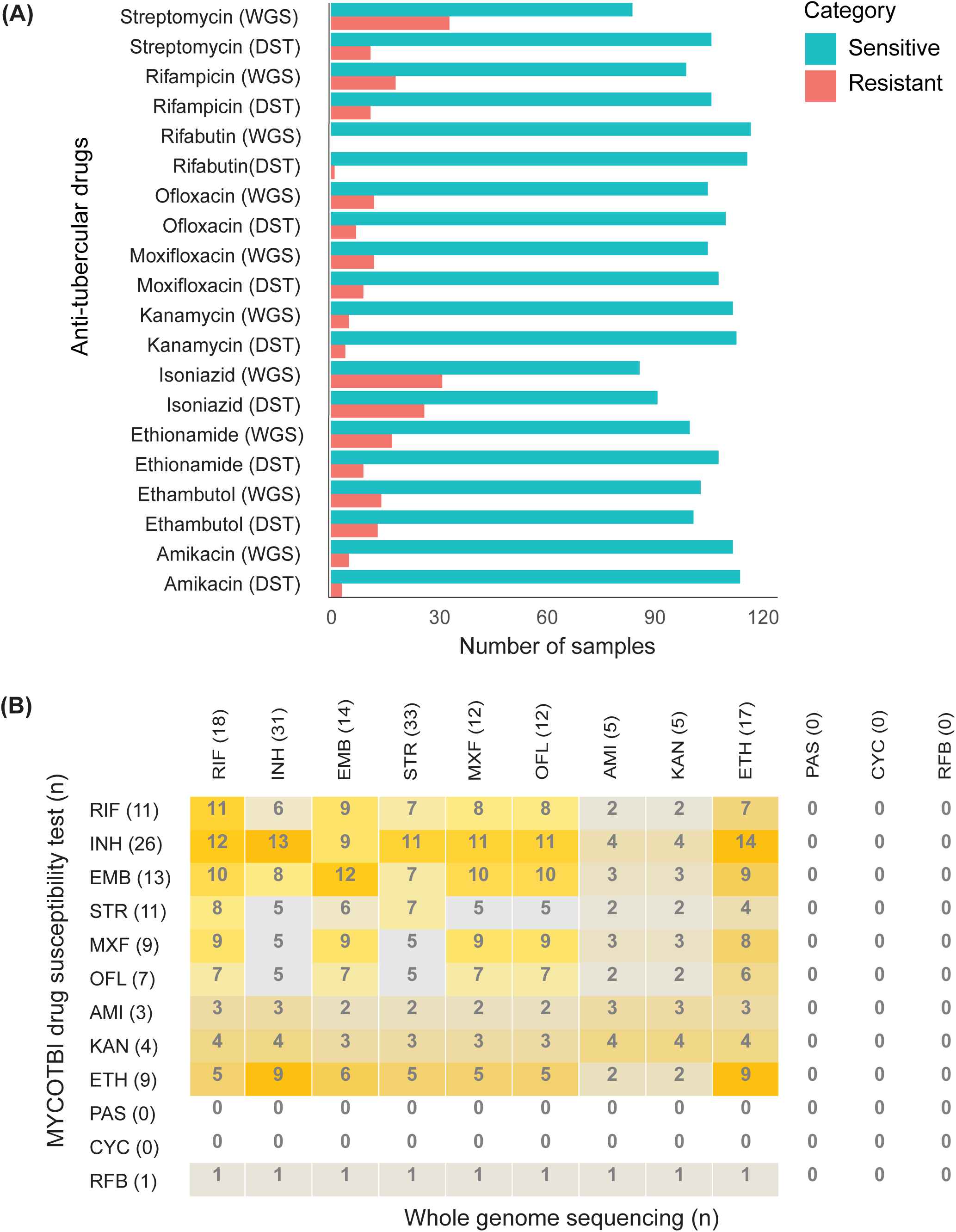
(A) MYCOTBI vs WGS resistance profile. A comparison of resistance and sensitivity profiles for twelve first-line and second-line anti-TB drugs was conducted using whole genome sequencing (WGS) and drug susceptibility testing (DST). Overall, 93% concordance was observed among the samples. Among the concordant cases (n = 90), 70 were drug-sensitive, 12 were isoniazid-monoresistant, 2 were rifampicin-monoresistant, 1 was MDR-TB, 3 were pre-XDR. **(B)** Concordance between whole genome sequencing (WGS) and drug susceptibility testing (DST) results for various anti-tuberculosis drugs. The numbers in parentheses indicate the total number of resistant samples for each drug. RIF – Rifampicin, INH – Isoniazid, EMB – Ethambutol, STR – Streptomycin, MXF - Moxifloxacin, OFL – Ofloxacin, AMI – Amikacin, KAN – Kanamycin, ETH – Ethionamide, PAS - Para-amino salicylic acid, CYC – Cycloserine, RFB – Rifabutin.

**Table 2B:**
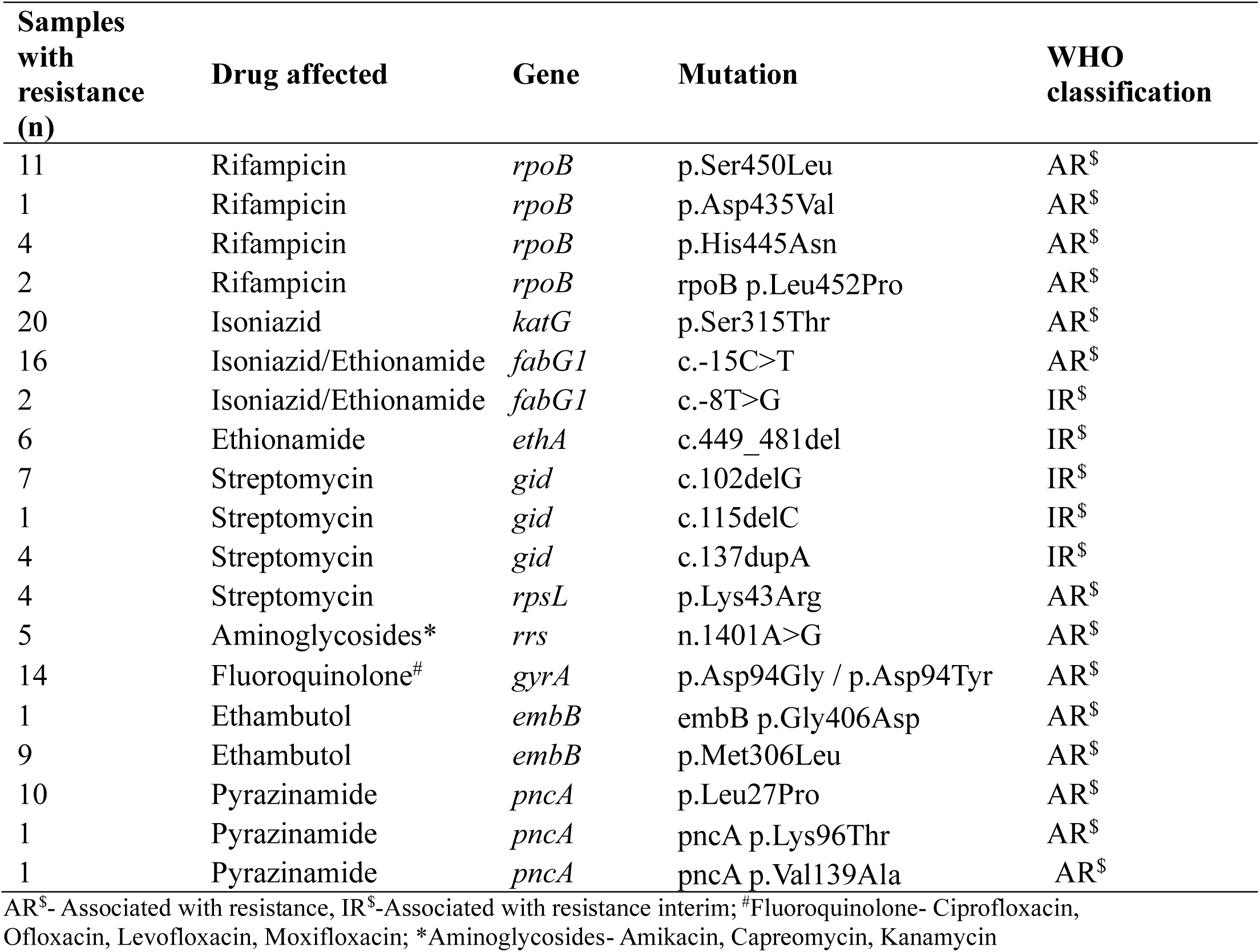
Drug resistance mutation profiles with the number of samples carrying mutations conferring drug resistance. The samples are classified according to the updated WHO catalog.

### Phenotype-genotype agreement

For the phenotype-to-genotype sensitivity analysis, we considered only those mutations listed in the WHO mutation catalogue as strongly associated with drug resistance. Based on this, WGS was selected as the reference standard to evaluate the sensitivity of the MYCOTBI drug DST for detecting resistance. A statistically significant difference in sensitivity between MYCOTBI DST and WGS was observed (McNemar’s p = 0.039). The MYCOTBI DST correctly identified 77.1% of resistant cases and 98.8% of sensitive cases as determined by WGS. These findings suggest that while MYCOTBI DST demonstrates high specificity (i.e., few false positives), it is less sensitive, missing approximately 23% of drug-resistant cases identified by WGS. Streptomycin exhibited the highest discordance between DST and WGS, with a difference observed in 22 cases. Additionally, two samples detected as rifampicin-sensitive by DST harbored *rpoB* p.His445Asn and *rpoB* p.Leu452Pro mutations, both known to confer high-level rifampicin resistance. Three additional samples carried the *rpoB* p.Ser450Leu mutation, a well-characterized resistance mutation included in the GeneXpert MTB/RIF assay. Furthermore, among the six discordant samples detected as isoniazid-sensitive by DST, four carried the *katG* p.Ser315Thr mutation, associated with high-level isoniazid resistance (**Figure 2B**) **(Figure 3)**. We found a significant association between sample type and resistance pattern (p = 0.0446). These findings suggest that the likelihood of detecting drug resistance may differ depending on the type of clinical sample.

**Figure 3:**
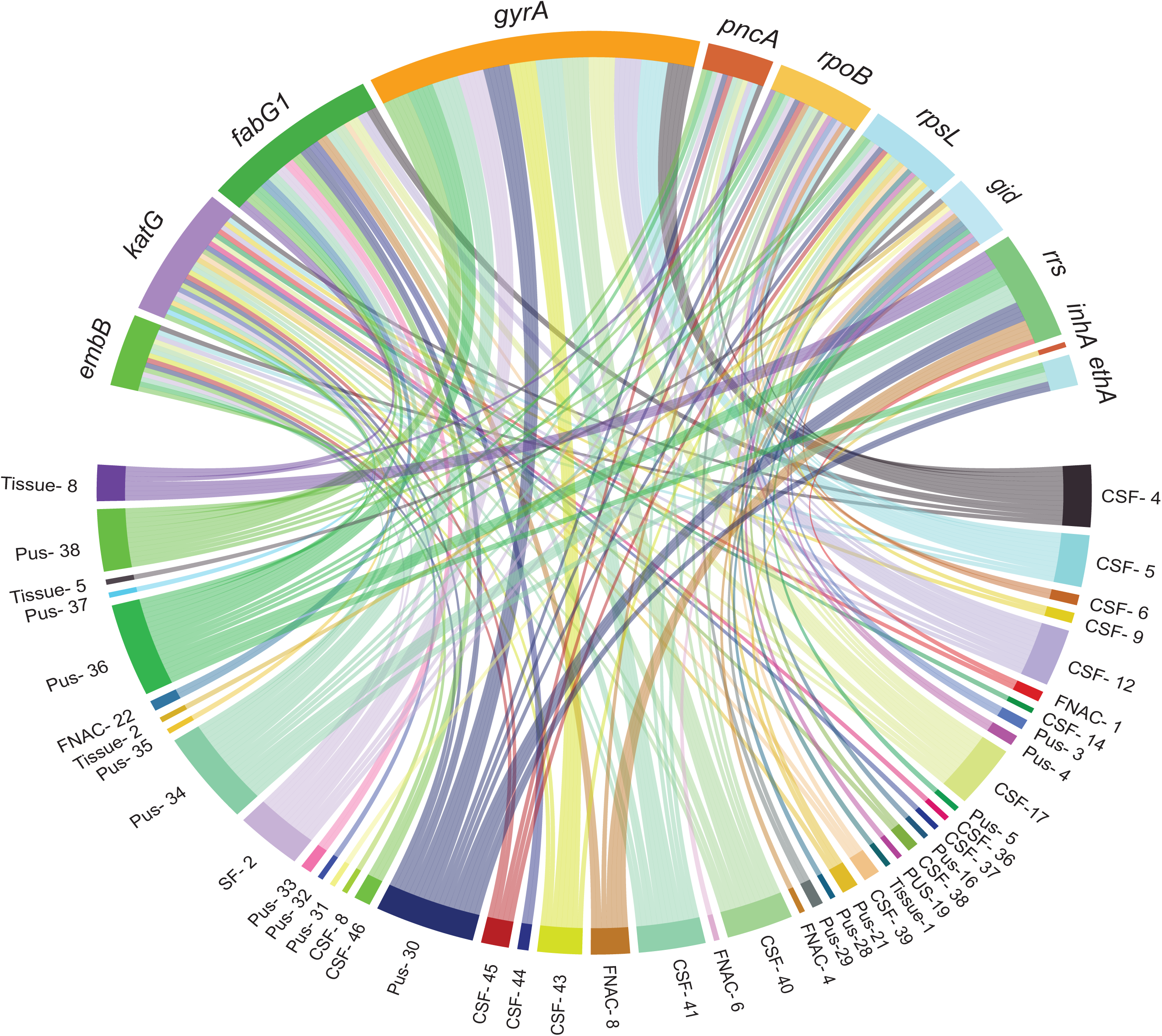
The Circos plot illustrates relationships between various genes and genomic regions associated with antibiotic resistance in *M. tuberculosis*. The upper half of the plot is labeled with gene names linked to resistance. Colored connections within the circle represent interactions or correlations between these genes and genomic regions. The dense network of lines underscores the complexity of genetic interactions and the multifactorial nature of antibiotic resistance.

### Lineage, clinical phenotype and drug resistance associations

Phylogenetic analysis successfully assigned lineages and sub-lineages to all 117 samples. Overall, four lineages (L1-L4) were detected in EPTB samples. Lineage (East-African Indian) which includes the Delhi/CAS (Central Asian Strain) family was the most predominant (73/117,62.3%), followed by lineage 2 (East Asian) (24/117, 23.3%), lineage 1 (Indo-Oceanic) (22/117, 20.9%), lineage 4 (7/117, 8.0%). These findings are in concordance with the previously reported prevalence of *M. tuberculosis* lineages from India [37] (**Figure 4**). After refining the dataset by excluding rare sample types and underrepresented lineages (<10), we observed a statistically significant association between sample type, lineage, and drug resistance (Chi-square p = 0.011). Lineage 2 exhibited a higher proportion of drug-resistant isolates (13/24, 54.1%), particularly from CSF samples (9/13, 69.2%), indicating a potential enrichment of resistant strains in extrapulmonary TB presentations involving the central nervous system. In contrast, when all sample types and lineages were included without filtering, the association did not reach statistical significance (p = 0.071).

**Figure 4:**
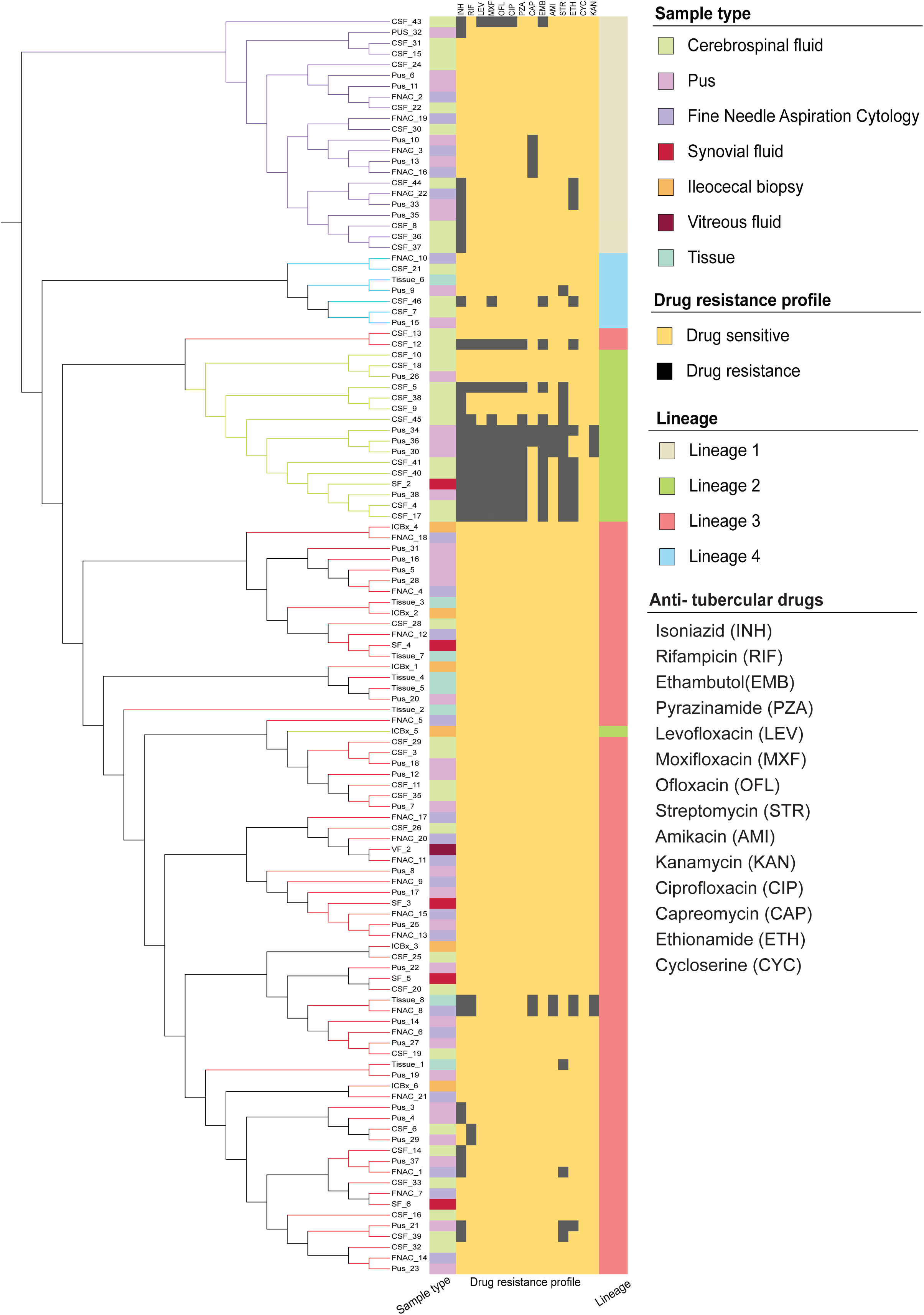
This phylogenetic tree represents the genetic lineages of the samples, categorized by sample type and their drug resistance profiles for anti-tuberculosis drugs. The adjacent heatmap shows drug resistance patterns, with yellow indicating drug-sensitive samples and black denoting drug-resistant samples.

### Mixed infections and heteroresistance

We observed mixed infections in 8/117 (6.8%) of the samples, the majority of which consisted of lineages 2 and 3 (7/8, 87.5%). One sample was infected with three lineages (L1, L2, L3). Lineage 4 was detected in 7/117 samples. Interestingly, we observed a high prevalence of drug resistance in lineage 2. At higher coverage depth, minority variants can be detected that may go undetected in phenotypic DST or other molecular methods. The presence of heteroresistance may correlate with the mixed infections and, in certain cases, explain the phenotype-genotype discordance. In our study, we detected heteroresistance in four samples: Pus_3, CSF_43, CSF_12, and CSF_8. **Table 3** contains the number of reads mapping to mutant and wild-type strains, along with the percentage of heteroresistance in each sample and the corresponding drug. Heteroresistance was significantly higher in mixed infections (p = 0.0139). Of the four samples with heteroresistance, two were infected with multiple strains (CSF_43 with lineages 1, 2, and 3, and CSF_10 with lineages 2 and 3). A lower number of resistant strains against a sensitive background may affect the positivity of resistance strains in culture. However, these are detected in WGS.

**Table 3:** Samples showing heteroresistance based on the reads mapped to both mutant and wild-type strains.

| **Sample ID** | **Reference nucleotide** | **Nucleotide position** | **Mutation nucleotide** | **Antibiotic** | **Total reads (Reference: Mutant)** |
| --- | --- | --- | --- | --- | --- |
| Pus_3 | C | 761139 | A | Rifampicin | 15:231 |
|  | C | 761155 | T | Rifampicin | 245:13 |
|  | A | 781687 | G | Streptomycin | 251:10 |
|  | A | 4247429 | G | Ethambutol | 297:10 |
|  | C | 2155168 | G | Isoniazid | 339:11 |
| CSF_12 | T | 2288955 | G | Pyrazinamide | 88:281 |
|  | A | 7582 | G | Fluoroquinolone | 77:211 |
|  | A | 4247429 | C | Ethambutol | 73:192 |
|  | C | 2155168 | G | Isoniazid | 108:289 |
|  | C | 1673425 | T | Isoniazid | 72:179 |
|  | A | 761110 | T | Rifampicin | 54:126 |
|  | A | 1472751 | G | Streptomycin | 73:125 |
| CSF_43 | A | 781687 | G | Streptomycin | 45:23 |
| CSF_8 | C | 4247863 | G | Ethambutol | 298:18 |

## DISCUSSION

Access to comprehensive drug resistance profiles in extrapulmonary tuberculosis is critical for guiding effective treatment regimens. Phenotypic DST is expensive and requires several weeks to provide results. Sequencing-based methods can reduce this turnaround time. However, current molecular and targeted sequencing approaches largely rely on genotype-phenotype associations derived from pulmonary TB, limiting their applicability to EPTB. Moreover, the tendency of specific *M. tuberculosis* lineages to localize in particular extrapulmonary sites, as well as the distribution of drug resistance across these lineages, remains poorly understood. In this study, we performed WGS of *M. tuberculosis* isolates cultured from seven distinct extrapulmonary sites and compared genotypic data with phenotypic DST results across twelve anti-TB drugs. We determined the phylogenetic lineage of each isolate and explored site-specific correlations between bacterial genotype and clinical phenotype. Additionally, we evaluated the prevalence of drug resistance across various EPTB sample types and investigated whether certain lineages or anatomical sites were associated with elevated resistance levels.

We observed drug resistance in 23.9% of isolates based on phenotypic DST and in 30.7% of samples using WGS. These findings are consistent with recently reported trends indicating a rise in drug resistance among EPTB cases [38,39]. We found high concordance (93%) between phenotypic and genotypic resistance profiles, aligning with previous studies reporting concordance rates ranging from 88.9% to 100% [40–42]. Notably, samples discordant with MYCOTBI DST results harbored mutations classified in the WHO mutation catalogue as conferring high-level resistance, suggesting true resistance and highlighting the superior sensitivity of WGS in detecting resistance-associated mutations [43,44]. Additionally, our study observed a higher frequency of drug resistance among EPTB isolates, corroborating prior reports [45]. There is also limited knowledge regarding the prevalence and clinical significance of mutations associated with resistance to newer anti-TB drugs in EPTB isolates, as these are underrepresented in the WHO mutation catalogue. To address this gap, we included four such drugs that included ofloxacin, cycloserine, PAS, and rifabutin in our analysis. While no phenotypic resistance was detected for these agents by the MYCOTBI assay, WGS revealed mutations associated with ofloxacin resistance in 12 samples and rifabutin resistance in 18 out of 117 samples. These findings underscore the need for expanded phenotype-to-genotype studies to better characterize resistance mutation patterns in EPTB and inform evidence-based treatment strategies.

Extensive scientific literature reports a high prevalence of resistance to first-line anti-tubercular drugs. This is likely due to their prolonged and widespread use in TB treatment. In contrast, second-line agents have generally been reserved for drug-resistant cases and are thus associated with lower resistance rates [45,46]. Given the higher frequency of resistance to first-line anti-tubercular drugs, it is essential to identify agents that remain effective in EPTB and exhibit lower rates of resistance-associated mutations. Prior studies have demonstrated that the pharmacokinetic properties of several anti-TB drugs hinder their penetration into extrapulmonary compartments. For example, two first line drugs rifampicin and ethambutol exhibit suboptimal CSF penetration [47]. The current WHO-recommended TB treatment regimen, which includes 8–12 mg/kg of rifampicin, CSF rifampicin levels often fall below the minimum inhibitory concentration required to effectively eliminate *M. tuberculosis* [2]. An increasing number of studies are exploring novel strategies that combine drugs with good penetration with high doses of drugs that typically have poor penetration, aiming to improve treatment outcomes. One study used linezolid and a higher dose of rifampicin to treat TB meningitis and reported optimal levels of linezolid in the CSF [48]. A clinical study reported significantly reduced mortality in TB meningitis patients treated with high-dose intravenous rifampicin (35%) compared to those receiving standard-dose therapy (65%) [11]. In contrast, fluoroquinolones show variable CSF penetration, with later-generation compounds such as levofloxacin and moxifloxacin achieving improved distribution [49,50]. Access to individualized drug resistance profiles can facilitate the design of personalized treatment regimens, potentially improving clinical outcomes.

As of current scientific consensus, at least nine major phylogenetic lineages of the *M. tuberculosis* have been identified, with lineages 1 through 4 being the most frequently encountered in global TB studies [51]. Despite this genetic diversity, current TB diagnostics and treatment regimens do not differentiate between pulmonary and EPTB with respect to lineage, nor do they account for its potential role in drug resistance. However, emerging studies suggest that certain *M. tuberculosis* lineages may be more prevalent in specific clinical presentations and associated with higher rates of drug resistance [13,52]. Higher drug resistance has been reported in Lineage 2 (Beijing strain) by several studies [13,53] A recent study reported that infection with Lineage 1 was more likely to result in TB osteomyelitis compared to other lineages [54]. In a large multinational study by Walker and colleagues involving 12,246 patients from three low-incidence and five high-incidence countries, pulmonary TB was significantly more likely in patients infected with lineage 2, 3, or 4 compared to lineage 1. Among pulmonary TB cases, Lineage 1 was associated with a higher risk of cavitary disease relative to Lineages 2 and 4. Conversely, in EPTB, Lineage 1 was more frequently linked to osteomyelitis compared to Lineages 2–4 [54]. In our study, we observed a significant predominance of Lineage 2 among isolates from CSF samples. Furthermore, we identified a statistically significant association between sample type, lineage, and drug resistance (p = 0.011). Lineage 2 exhibited a high proportion of drug-resistant isolates (13/17, 76.5%), particularly among CSF samples (13/23, 56.5%). These findings support the hypothesis of lineage-specific enrichment in particular anatomical compartments. Further studies with larger sample sizes are needed to validate these associations and assess their clinical implications.

Another underexplored factor that may influence the prevalence of drug resistance is the presence of multiple *M. tuberculosis* strains within a single clinical sample. Co-infection with more than one strain, including drug-resistant variants, has been documented in TB patients [55]. When resistant strains are present in a minority, they may go undetected by conventional DST, as dominant drug-sensitive strains can outgrow them during culture. Detecting such minority variants is critical for guiding appropriate treatment regimens.

In our study, we developed a WGS-based pipeline capable of detecting minority strains and resistance-associated mutations within mixed infections. We found that heteroresistance was significantly more frequent in samples with mixed strain infections (p = 0.0139). These findings suggest that mixed infections substantially increase the likelihood of heteroresistance, which may compromise treatment outcomes. Our results underscore the need to incorporate lineage-specific markers in routine diagnostic workflows to improve the detection and management of complex TB infections.

The findings of our study are subject to several limitations. First, we sequenced a very small number of samples, including tissue (n = 8), ileocaecal biopsy (n = 6), synovial fluid (n = 5), and vitreous fluid (n = 1). Further studies with larger sample sizes across these categories are needed to validate whether certain lineages preferentially colonize these sample types. Additionally, our analyses were based on cultured isolates, which may have reduced the observed genetic diversity and led to missed cases of mixed infections. Although we detected mixed infections in eight samples, and these were significantly correlated with heteroresistance, larger studies are needed to confirm these findings.

This study demonstrates that WGS is a highly effective tool for detecting drug resistance in EPTB, exhibiting a high phenotype-to-genotype concordance of 93.5%. WGS enables the identification of mutations associated with high-level resistance, even in samples that appear phenotypically susceptible, and can detect minority variants often missed by conventional DST. Furthermore, our findings suggest a preferential distribution of certain *M. tuberculosis* lineages based on clinical phenotype, with some lineages exhibiting a higher frequency of drug resistance. WGS thus offers a reliable alternative to traditional DST, especially in cases of low culture positivity and limited drug penetration at extrapulmonary sites. These insights highlight the potential of WGS to enhance drug resistance detection and inform more precise, effective treatment strategies for EPTB.

## Data Availability

All data produced in the present study are available upon reasonable request to the authors

## Author Contributions

KS, RV, and AP conceptualized and designed the study. KS, AS, and MM attended patients and provided clinical data. RV and KS analyzed and interpreted the data and wrote the manuscript. AB processed the samples, performed DNA extraction, and contributed to data analysis. GKM, MR, KV, and JG performed WGS analysis and prepared the figures. ASB and PP contributed to data analysis, figures, and tables. SV and PY also worked on data analysis, tables, and figures. KV and JS supervised the computational team. AG, SKS, MSD, VS, RS, and NS collected clinical samples and done patient follow-up. MS has done data curation, and sample processing. RN did histopathological examination of tissue samples. KS and AP provided input during data analysis and interpretation. All authors reviewed the final manuscript and provided their feedback.

## Acknowledgements

The authors acknowledge Dr. Neeraj Singla, Dr. Ritu Shree, Dr. Ashish Kumar Kakkar, Dr. Apinderpreet Singh, Dr. Sameer Vyas, Dr. Chirag Kamal Ahuja, Dr. Vijeta Patial, Dr. Riya Sharma, Dr. Siddharth Chand, Dr. Yatharth Dixit, Dr. Himanshu, Pratibha Chauhan and all of the technical staff and patients from PGIMER Hospital for their cooperation during the study.

## Data availability

Data supporting the findings of this manuscript have been submitted to NCBI and can be accessed through the Sequence Read Archive submission portal with accession number PRJNA1273668 (https://www.ncbi.nlm.nih.gov/sra/PRJNA1273668).

